# Solar-powered phototherapy for severe neonatal hyperbilirubinemia in low-resource settings: a multicentre implementation study in Nigeria

**DOI:** 10.64898/2026.02.04.26345610

**Authors:** Hippolite O. Amadi, Obot E. Antia-Obong, Leo-Stanley Chinonso Muoneke, Martin M. Meremikwu, Jacob J. Udo, Sarah Ango

**Affiliations:** Department of Bioengineering, Imperial College London, United Kingdom; Department of Paediatrics, University of Calabar Teaching Hospital, Nigeria; Department of Paediatrics, Federal Medical Centre, Keffi, Nigeria

**Keywords:** Neonatal hyperbilirubinemia, Solar-powered phototherapy, Implementation research, Task shifting, Low- and middle-income countries, Climate-resilient healthcare

## Abstract

Severe neonatal hyperbilirubinemia remains a leading cause of preventable neonatal morbidity and mortality in low- and middle-income countries (LMICs), where unreliable electricity supply, limited specialist availability, and high treatment costs constrain access to effective phototherapy. There is limited evidence on scalable, climate-resilient phototherapy solutions suitable for decentralised care delivery.

We conducted a multicentre implementation study evaluating a fully solar-powered total-body phototherapy system deployed across tertiary and grassroots healthcare facilities in Nigeria between February 2024 and January 2026. The study followed a phased design: (1) specialist-controlled observational deployment at a tertiary hospital, (2) corroborative deployment at additional tertiary centres across climatic regions, and (3) task-shifted grassroots implementation in mission-owned hospitals. Neonates with moderate-to-severe hyperbilirubinemia were treated according to standard clinical protocols. Primary outcomes were successful reduction of bilirubin to ≤11.5 mg/dL, treatment duration, safety, and operational feasibility under electricity-independent conditions.

Across all phases, nearly 2000 neonates have been screened and more than 650 neonates received treatment with the solar-powered phototherapy system. All treated neonates achieved bilirubin reduction to the predefined safety threshold, with a mean treatment duration of 19.2 ± 13 hours during the initial observational phase. No treatment interruptions due to power failure occurred, and no phototherapy-related adverse events, neurological complications, or treatment-related mortality were observed. Treatment was safely delivered by junior clinicians and health assistants under task-shifted models in grassroots settings. The intervention eliminated the need for exchange blood transfusion among treated neonates and substantially reduced hospitalisation duration compared with conventional care pathways.

This study demonstrates that fully solar-powered phototherapy can be safely, effectively, and sustainably implemented across multiple levels of the health system in a low-resource setting. By removing dependence on grid electricity and specialist personnel, the intervention addresses key structural barriers to neonatal care while supporting task shifting and climate-resilient service delivery. The findings support policy consideration of decentralised, infrastructure-independent phototherapy as a strategy to reduce neonatal morbidity and mortality in LMICs.

## Introduction

Severe neonatal jaundice (SNJ) remains a major but preventable contributor to neonatal morbidity and mortality in low- and middle-income countries (LMICs), particularly in sub-Saharan Africa [1–3]. Nigeria bears a disproportionate share of this burden, with high incidence, late presentation, and limited access to timely and effective treatment contributing to avoidable deaths and long-term neurodevelopmental impairment [4,5]. Despite decades of clinical awareness and policy attention, population-level outcomes have improved only marginally, underscoring persistent structural and health-system constraints, particularly unreliable power [6,7].

Conventional management of SNJ relies on grid-powered phototherapy and, for severe cases, exchange blood transfusion (EBT). In LMIC settings, both modalities are frequently compromised by unreliable electricity supply, shortages of trained personnel, delayed referrals, and prohibitive out-of-pocket costs [8–11]. In Nigeria, chronic electricity instability remains a systemic challenge, forcing health facilities to depend on diesel generators or to ration phototherapy use, often resulting in interrupted treatment or complete service unavailability [12–14]. These constraints contribute directly to the continued use of EBT for bilirubin levels that would otherwise be amenable to intensive phototherapy, exposing neonates to avoidable procedural risks and prolonged hospitalisation [15,16].

Recent implementation and health-systems research have highlighted the importance of context-compatible technologies and task-shifting strategies to address care gaps in resource-limited settings [17–19]. In neonatal care, innovations that reduce dependence on continuous electricity supply, minimise the need for highly specialised staff, and lower treatment costs are particularly critical for extending life-saving interventions to rural and peri-urban populations [20,21].

Evidence from Nigeria and comparable LMICs suggests that decentralised, non-invasive interventions could substantially reduce delays to care and prevent progression to severe hyperbilirubinemia [19,22,23].

Total-body phototherapy has previously been shown to improve bilirubin clearance rates and reduce treatment duration compared with conventional overhead phototherapy in Nigerian clinical settings [16,21]. However, these systems have largely remained tethered to the same infrastructural vulnerabilities that limit conventional devices. The development of a fully solar-powered, self-sustaining phototherapy platform therefore represents a potentially transformative advance, particularly for settings where grid electricity and specialist paediatric care are unreliable or absent.

This study evaluates the phased implementation of a novel solar-powered total-body phototherapy device (PUL) for the treatment of neonatal hyperbilirubinemia in Nigeria. Using a staged implementation approach—comprising specialist-controlled deployment at a tertiary centre, corroborative evaluation across additional referral hospitals, and subsequent rollout to grassroots facilities—we assess feasibility, clinical performance, safety, and early health-system implications. By situating this intervention within the broader challenges of neonatal care delivery, energy insecurity, and climate-conscious health innovation, this work aims to inform scalable strategies for reducing SNJ-related morbidity and mortality in Nigeria and other LMICs.

## Materials and Methods

### Study design and reporting framework

This study was a multicentre, staged implementation study evaluating a fully solar-powered phototherapy system (PoliteUltralumen; PUL) for the treatment of severe neonatal hyperbilirubinemia in Nigeria (Fig. 1). The work followed an observational design with prospective data capture across sequential deployment phases and is reported in accordance with the Strengthening the Reporting of Observational Studies in Epidemiology (STROBE) guidelines [14,24].

**Figure 1:**
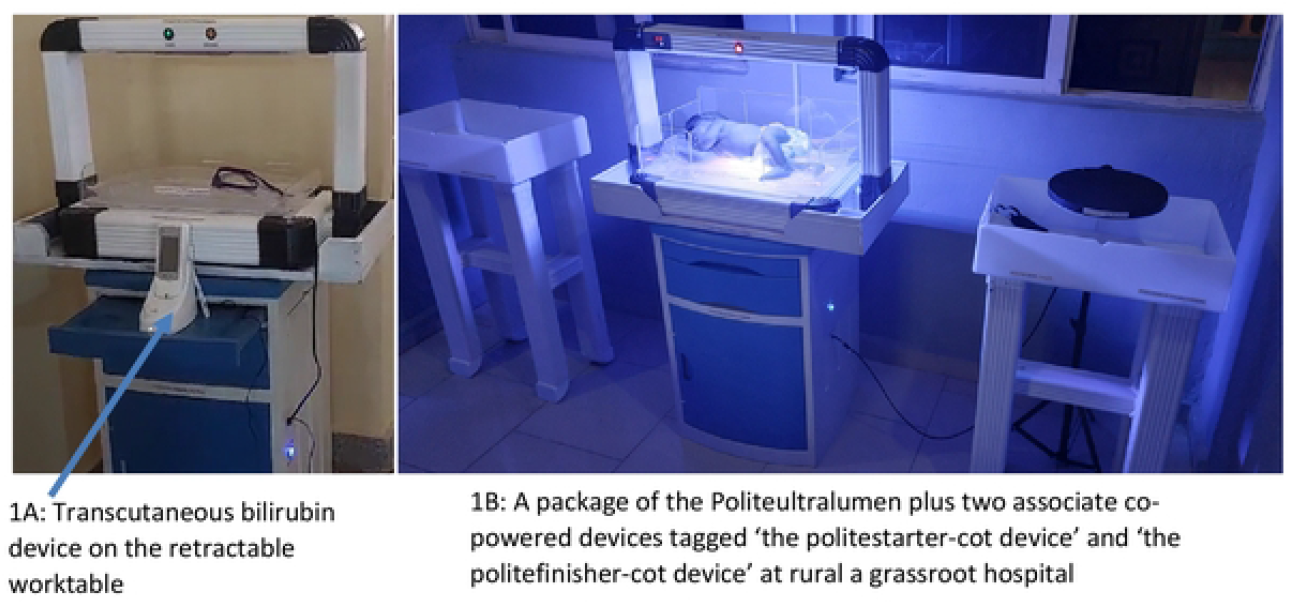
The politeultralumen device with a transcutaneous bilirubinometer.

### Study setting and phased implementation

The study was conducted across tertiary and grassroots healthcare facilities in Nigeria, a setting characterised by high neonatal jaundice burden and persistent electricity unreliability [3,4,6,7]. Implementation occurred in three predefined stages designed to progressively assess safety, feasibility, and scalability under real-world constraints (Fig. 2).

**Figure 2:**
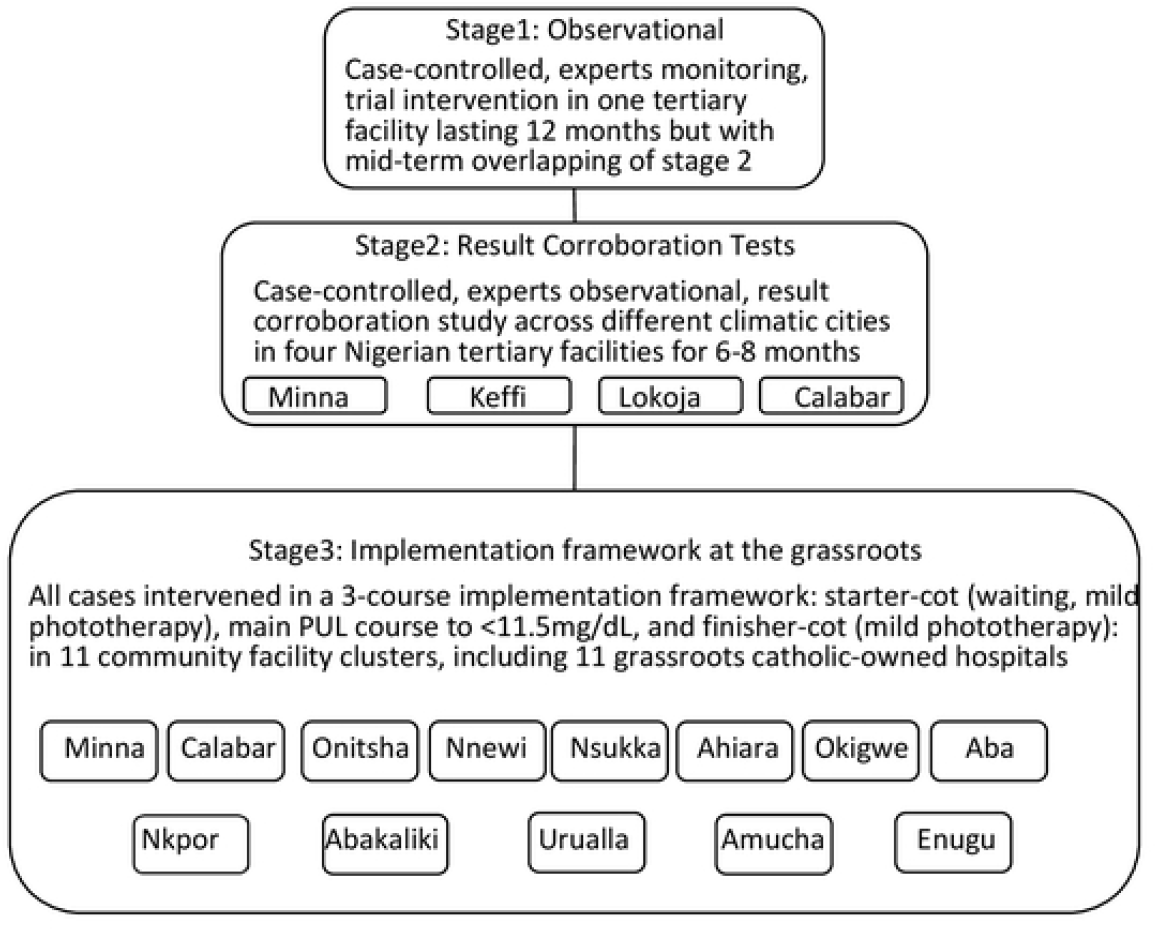
Flowchart of the 3-staged study design.

### Stage 1 (specialist-controlled implementation)

Initial deployment occurred at the Special Care Baby Unit (SCBU) of the University of Calabar Teaching Hospital (UCTH) Nigeria in January 2024. During this phase, use of the PUL device was supervised by paediatric specialists to establish safety, operational stability, and early clinical performance under controlled conditions. Only one device unit was available at this site, necessitating sequential patient access.

### Stage 2 (tertiary-centre corroboration)

Following successful initial operation, the device was deployed to multiple tertiary hospitals across different geographic and climatic regions of Nigeria. This phase focused on corroborating performance consistency, operational resilience, and early implementation challenges at scale, particularly within referral centres managing high neonatal volumes.

### Stage 3 (grassroots and task-shifted implementation)

Subsequent deployment occurred at selected grassroots and primary-level facilities to evaluate feasibility under task-shifted care models [25–27]. In this phase, treatment was delivered primarily by junior doctors, nurses, and health assistants without continuous on-site consultant supervision, reflecting real-world human-resource constraints common in low- and middle-income countries (LMICs) [16,21].

### Participants and eligibility criteria

Neonates admitted to participating facilities with clinically significant hyperbilirubinemia requiring phototherapy were eligible for inclusion. Inclusion criteria comprised gestational age ≥30 weeks, birthweight ≥1.5 kg, and elevated bilirubin levels above locally defined treatment thresholds consistent with Nigerian clinical practice [3,4,21]. Patient carers were mandatorily made to sign the consent form after a thorough explanation of the new intervention technique and their acceptance of inclusion. Neonates requiring immediate exchange blood transfusion (EBT) at presentation or presenting with major congenital anomalies incompatible with phototherapy were excluded.

### Intervention: solar-powered total-body phototherapy

The PUL device is a fully self-powered phototherapy system incorporating solar energy capture, battery storage, and dual-surface total-body irradiation. The system delivers continuous blue-light irradiation from both overhead and base lamps, eliminating reliance on grid electricity and mitigating treatment interruption due to power outages—a major limitation of conventional phototherapy in Nigeria [4,6,7].

Irradiance levels were monitored during treatment at standardised anatomical points (trunk, cranial/caudal, and lateral approximations), using a calibrated light irradiation meter (MTTS Light Meter, Medical Technology Transfer and Services Co. Ltd., Hanoi, Vietnam). Treatment was paused only for breastfeeding and routine neonatal care.

### Clinical procedures and monitoring

Baseline demographic and clinical data were collected at admission, including gestational age, birthweight, mode and place of delivery, and bilirubin levels. Total serum bilirubin (TSB) or transcutaneous bilirubin (TcB) measurements were obtained at baseline and serially during treatment using validated bilirubinometry techniques appropriate for low-resource settings [17,22,28].

Phototherapy was continued until bilirubin levels declined to a predefined mild benchmark (≤11.5 mg/dL), consistent with prior Nigerian phototherapy outcome studies [4,9]. Neonates were continuously monitored for vital signs, hydration status, and potential adverse events, including dermatological, ophthalmological, neurological, or audiological complications [23,24].

### Outcomes

The primary outcome was successful bilirubin reduction to the predefined mild benchmark without the need for EBT. Secondary outcomes included rate of bilirubin

decline (mg/dL/hour), treatment duration, occurrence of adverse events, need for escalation to invasive interventions, and operational continuity of the device (Fig. 3).

**Figure 3:**
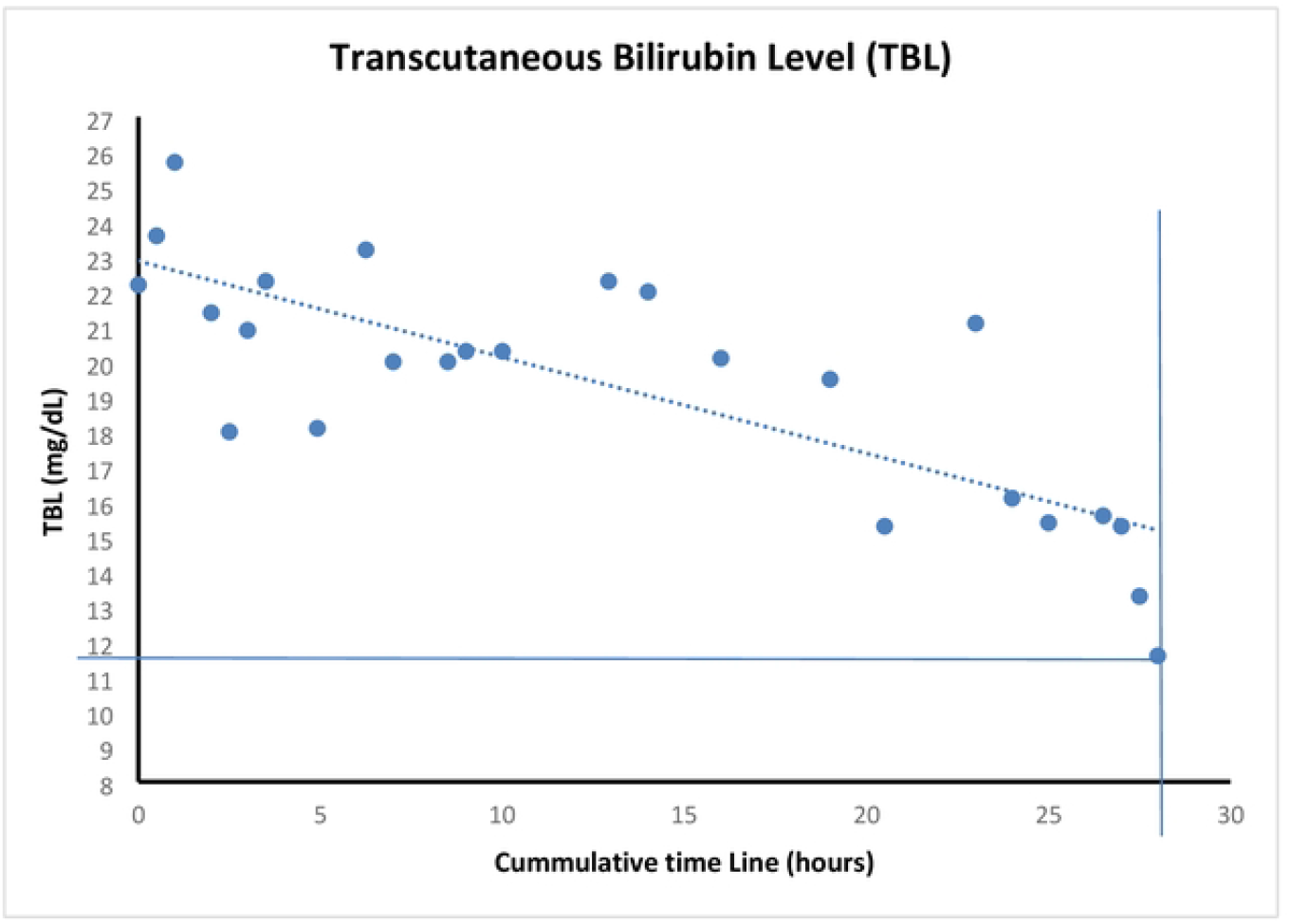
A typical plot showing the degradation of transcutaneous bilirubin level (TBL) during treatment.

Implementation outcomes included system uptime, irradiation stability, need for maintenance or repairs, and feasibility of use under task-shifted staffing models [16−18,27].

### Follow-up

All treated neonates underwent routine clinical follow-up according to institutional protocols. Specialist assessments were conducted where available to screen for delayed dermatological, ophthalmological, neurological, or audiological sequelae associated with severe hyperbilirubinemia or phototherapy exposure [16,23].

### Ethical considerations

Ethical approval was obtained from the relevant institutional review boards overseeing participating centres. Written informed consent was obtained from parents or legal guardians prior to treatment. The study adhered to the principles of the Declaration of Helsinki and applicable Nigerian research governance regulations.

### Data management and analysis

Data were prospectively recorded using standardised case-record forms and anonymised prior to analysis. Descriptive statistics were used to summarise demographic characteristics, bilirubin trajectories, treatment duration, and outcomes. Continuous variables are presented as ranges, means, and standard deviations, while categorical variables are presented as frequencies and proportions. Given the implementation-focused design, no formal hypothesis testing was undertaken.

## Results

### Patient flow and baseline characteristics

During the specialist-controlled implementation phase at UCTH Calabar (Stage 1), a total of 218 neonates with clinically significant hyperbilirubinemia presented to the Special Care Baby Unit between February 2024 and February 2025. Of these, 79 (36.2%) were inborn and 139 (63.8%) were referred from external facilities. Due to the availability of a single PUL unit, 42 neonates (19.3%) received treatment with the device during this initial phase.

Baseline characteristics of treated neonates included birthweights ranging from 1.5 to 4.3 kg, gestational ages of 30–41 weeks, and peak pre-treatment bilirubin levels of 12.6–25.7 mg/dL. These characteristics were consistent with locally defined thresholds for severe neonatal hyperbilirubinemia requiring intervention.

### Treatment delivery and device performance

Across all treated cases in Stage 1, phototherapy was delivered without interruption once initiated. No device-related power outages occurred, and continuous irradiation was maintained throughout each treatment course, except for brief pauses during breastfeeding and routine neonatal care.

Irradiance measurements obtained at trunk, cranial/caudal, and lateral anatomical points demonstrated stable output from both the overhead and base lamps throughout treatment periods (Fig. 4). No clinically meaningful reduction in irradiation intensity was observed during evening or sunset hours. No system failure, repair, or maintenance callout was required at UCTH Calabar during the first 24 months following installation.

**Figure 4:**
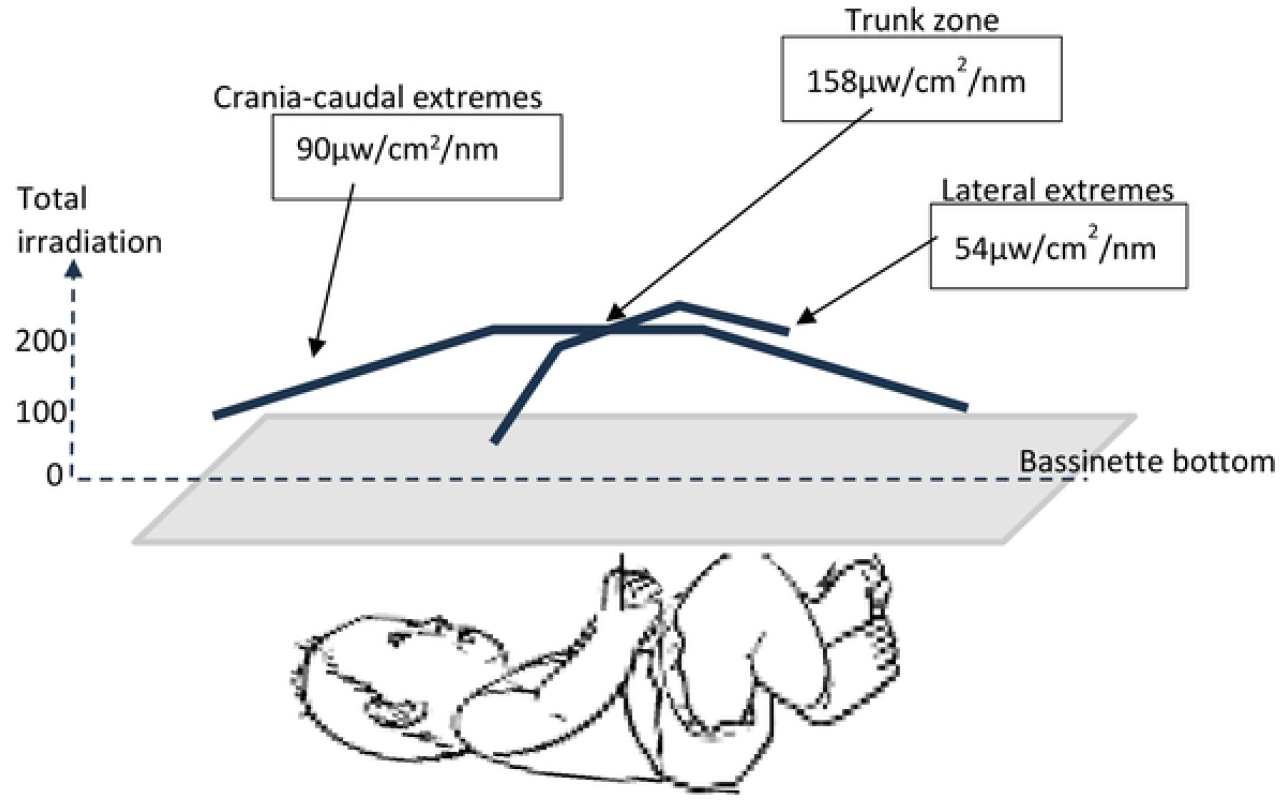
Irradiation distribution across the body segments of a neonate.

### Clinical outcomes (Stage 1)

All 42 neonates treated with the PUL device during Stage 1 achieved successful bilirubin reduction to the predefined mild benchmark (≤11.5 mg/dL). The fractional reduction in transcutaneous bilirubin from peak value to treatment cessation ranged from 9% to 57%.

Mean treatment duration was 19.2 ± 13 hours. The average rate of bilirubin decline was 0.3 mg/dL per hour. No neonate required escalation to exchange blood transfusion or adjunctive phototherapy modalities (Table 1).

**Table 1.** Clinical outcomes stratified by baseline hyperbilirubinemia severity. Clinical characteristics and treatment outcomes of neonates treated with the politeultralumen (PUL) device during the specialist-controlled observational phase at UCTH Calabar. Severity groups are defined by peak pre-treatment transcutaneous bilirubin level (TBL). Outcomes include treatment duration, rate of bilirubin decline, and achievement of the predefined treatment endpoint (TBL ≤11.5 mg/dL).

| Patient TSB group | No. | Ave BW (kg) | Average GA (weeks) | Ave Peak TBL (mg/dL) | Ave End TBL (mg/dL) | Ave TBL drop (%) | Ave Duration (hr) | Treatment speed (mg/dL/hr) | Overall Outcome |
| --- | --- | --- | --- | --- | --- | --- | --- | --- | --- |
| **Overall** | **42** | **3.0** | **37.6** | **16.2** | **11.2** | **28.8** | **19.2** | **0.3** | **Discharge without issues or negative events** |
| **SD** |  | **0.6** | **2.4** | **3.1** | **0.5** | **12.1** | **12.9** | **0.2** |  |
| 12-13.9 | 11 | 2.6 | 37.2 | 13.3 | 11.4 | 14.4 | 8.6 | 0.3 |  |
| 14-16.9 | 19 | 3.0 | 38.1 | 15.5 | 11.1 | 27.9 | 21.8 | 0.3 |  |
| 17-19.9 | 8 | 3.1 | 37.0 | 18.1 | 11.1 | 38.8 | 21.8 | 0.4 |  |
| 20-21.9 | 1 |  |  | 20.9 | 11.2 | 46.4 | 23.5 | 0.4 |  |
| 22-24.9 | 1 | 1.9 | 34.0 | 23.2 | 11.2 | 51.7 | 59.5 | 0.2 |  |
| >25 | 2 | 3.1 | 40.0 | 25.6 | 11.3 | 55.8 | 21.0 | 0.8 |  |

*Note.* TSB. = total serum bilirubin; TBL. = transcutaneous bilirubin level; BW. = birthweight; GA. = gestational age

No abnormalities in vital signs were observed during treatment. There were no reported dermatological or ophthalmological complications, and no cases of acute neurological or audiological concern were identified during treatment or follow-up assessments.

### Outcomes by severity category

Treatment outcomes stratified by pre-treatment bilirubin severity demonstrated consistent effectiveness across severity groups. Neonates with higher baseline bilirubin levels required longer treatment durations but achieved comparable bilirubin clearance without adverse events. Severity-stratified outcomes are summarised in Table 1.

### Multicentre corroboration and task-shifted implementation (Stages 2 and 3)

During Stage 2, the PUL device was deployed to multiple tertiary hospitals across Nigeria for corroborative implementation. All neonates treated during this phase were successfully discharged following bilirubin reduction without escalation to invasive interventions.

However, all tertiary corroboration centres reported mechanical failure of the patient bassinette associated with a specific manufacturing batch used during this phase.

These failures did not result in reported clinical harm but led to temporary suspension of device use at some sites prior to replacement and redesign. No additional device-related clinical incidents were reported.

Stage 3 deployment occurred at selected grassroots and primary-level facilities using redesigned hardware. Treatment was delivered primarily by junior doctors, nurses, and health assistants under task-shifted care models. All neonates treated during this phase to date were discharged successfully and no post-treatment adverse issue has been raised from any of the 11 centres. At the time of manuscript preparation, follow-up duration for Stage 3 remained insufficient for definitive impact assessment; extended evaluation is ongoing.

### Aggregate outcomes across all stages

By the time of data analysis, more than 650 neonates had been treated with the PUL device across all stages and participating regions in Nigeria with nearly 2000 neonates benefiting from the diagnostic community screening exercise. No treatment failures, mortality, or serious adverse clinical events were reported among treated neonates (Table 2).

**Table 2.**
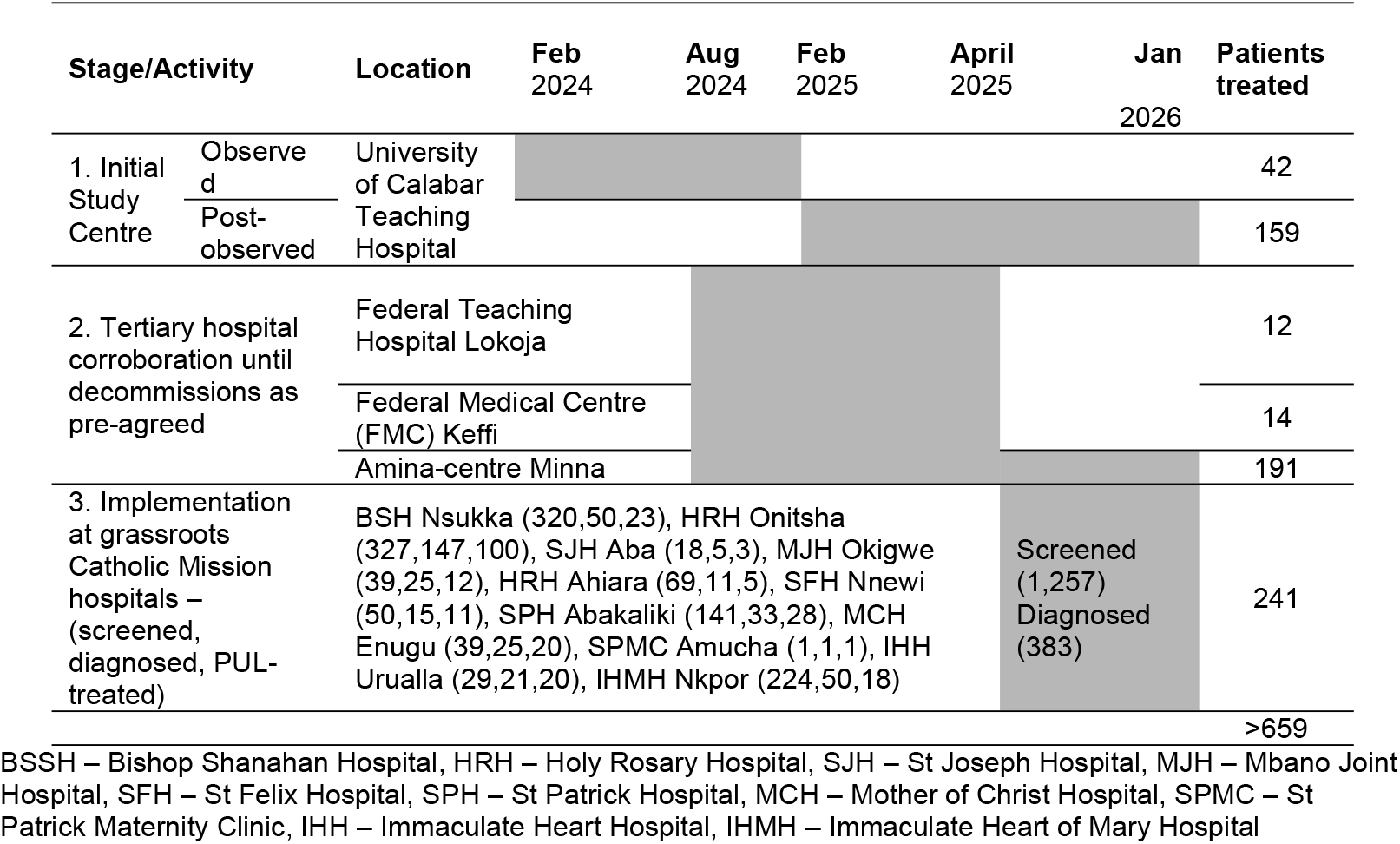
Timeline to specialist-controlled observational (Stage 1), tertiary corroboration across climatic regions of Nigeria (Stage 2), and implementation at grassroots level (Stage 3). Follow-up for stage 3 remains ongoing at the time of reporting.

## Discussion

This study demonstrates that a fully solar-powered total-body phototherapy system can be safely and effectively implemented across multiple levels of the health system in a low-resource setting, achieving complete treatment success without reliance on grid electricity or specialist-intensive care. The findings address long-standing structural barriers to neonatal care in LMICs, where infrastructure instability, workforce shortages, and household poverty continue to drive preventable neonatal morbidity and mortality [1,3,4].

### Implementation relevance and health system impact

The intervention was initiated in response to a pandemic-scale burden of severe neonatal hyperbilirubinemia within a tertiary facility and subsequently expanded through deliberate phases of corroboration and grassroots implementation. This phased approach allowed real-world assessment of feasibility, safety, and adaptability across heterogeneous clinical and climatic contexts, a core requirement of implementation research [26]. Importantly, the system functioned without interruption over extended periods, underscoring the operational resilience required for decentralised neonatal care in electricity-insecure settings [6–8].

The observed elimination of exchange blood transfusion among treated neonates is particularly relevant in the Nigerian context, where exchange transfusion remains common for severe hyperbilirubinemia despite its invasive nature, procedural risks, and high resource requirements [10,16,23]. By enabling rapid bilirubin reduction using non-invasive phototherapy, the intervention supports safer care pathways and reduces dependence on high-risk procedures.

### Task shifting and workforce considerations

A key finding of this study is the safe delivery of phototherapy by junior clinicians, nurses, and health assistants under supervision, including during the grassroots implementation phase. This aligns with growing evidence that task shifting is an effective strategy for addressing workforce shortages in LMIC health systems when supported by appropriate technology and protocols [17,18,27]. The ability to deliver effective treatment without on-site specialist presence has important implications for rural and mission-owned facilities, where paediatric expertise is often unavailable.

The results extend prior task-shifting literature by demonstrating that technology design itself—specifically electricity independence, simplified operation, and continuous functionality—can actively enable safe delegation of care tasks rather than merely reallocating existing clinical responsibilities [17,18].

### Climate resilience and sustainability

The solar-powered design of the phototherapy system positions it within the emerging discourse on climate-resilient healthcare. Health systems in LMICs are disproportionately vulnerable to climate-related disruptions, including power instability and fuel insecurity [25]. By operating independently of fossil fuels and grid electricity, the intervention reduces both service fragility and the carbon footprint associated with neonatal care, including generator use and long-distance patient transfers to urban centres.

This dual benefit—health system strengthening and climate mitigation—supports calls for context-specific, low-carbon innovations as part of national adaptation strategies for health [25].

#### Cost implications and equity

Although formal economic evaluation was not the primary focus of this study, the substantial reduction in hospitalisation duration and elimination of electricity and exchange transfusion costs are consistent with previous findings demonstrating major cost savings from total-body phototherapy approaches [16,21]. When extended to rural settings, these savings may be amplified by reduced travel,accommodation, and opportunity costs borne by families—factors known to contribute to delayed or foregone care and adverse neonatal outcomes.

By lowering both system-level and household-level costs, the intervention directly advances equity in access to life-saving neonatal care.

### Limitations

This study has limitations. Meteorological instrumentation to formally quantify the relationship between weather variability and device performance was not available during the initial phases but is planned for future work. Hardware failures affecting the patient bassinette were reported at some corroborating centres during the second phase of implementation, necessitating redesign and temporary service interruption. While these issues did not result in clinical harm, they reduced patient throughput during that phase and highlight the importance of iterative hardware refinement during scale-up.

In addition, long-term neurodevelopmental follow-up is ongoing, and future analyses will be required to confirm sustained outcomes beyond early infancy.

### Policy implications

The findings support policy consideration of decentralised, electricity-independent phototherapy as part of national newborn care strategies in LMICs. Integrating such technologies into primary and secondary care platforms could reduce referral bottlenecks, lower neonatal mortality, and enhance system resilience in the face of infrastructure and climate constraints [28,29]. More broadly, this study illustrates the potential of locally driven innovation to generate scalable solutions to entrenched global health challenges [26,29].

## Data availability

De-identified data generated during this study will be deposited in the Imperial College London institutional repository and made publicly available upon publication.

## Acknowledgements

This project has indirectly been supported by the Nigeria Liquefied Natural Gas (NLNG) Limited unknown to them as the financial reward from Hippolite’s win of the Nigeria Prize for Science, 2023 (https://www.nigerialng.com/csr/Pages/The-Nigeria-Prizes.aspx), sponsored by the NLNG was the main source of funds for the PUL research, development and clinical trialling. The medical missions of Professor H Amadi in Nigeria is anchored by Neonatal Concerns for Africa (www.neonatalconcerns.org)—a collaboration of the Bioengineering Department of Imperial College London, and in part, supported by the Hornchurch Baptist Church, Essex England, United Kingdom (https://www.hornchurchbaptist.org.uk). We acknowledge all technical supports and assistance received from Neonatal Concerns for Africa technical team. We appreciate the kind collaborative support of the Catholic bishops and dioceses whose grassroots hospitals participated in Stage 3 of this project across the southeast Nigeria. The Institute of Tropical Disease Research and Prevention of the University of Calabar provided their staff assistants for patient monitoring during treatment and supports for diagnostics and laboratory tests at Stage 1.

